# Mortality from birth through adolescence: Trends, Determinants, and Insights from a Longitudinal Cohort in Benin

**DOI:** 10.64898/2026.06.23.26356407

**Authors:** Katharine M. Barry, Melissa Cruz, Maroufou Jules Alao, Romeo Zoumenou, Christophe Fermanian, Achille Massougbodji, Florence Bodeau-Livinec

## Abstract

**Introduction:** Despite substantial global progress, child mortality remains a major public health burden, with sub-Saharan Africa disproportionately affected. In West Africa, deaths from preventable causes remain alarmingly high, yet longitudinal evidence on the rates and risk factors of child mortality in the region is scarce. This study is among the first in Benin to examine child mortality from birth to 14 years in a community-based, longitudinal cohort.

**Methods:** Data originate from the Malaria in Pregnancy Preventive Alternative Drugs (MiPPAD) trial (NCT00811421), an open-label, randomized controlled trial that recruited pregnant women in their second trimester across four sub-Saharan African countries between September 2009 and December 2012. This analysis uses data from 1183 women enrolled at three health centers in the semi-rural Allada district of Benin, with offspring follow-up at 1, 9, and 12 months, 6 years, and 13–14 years.

**Results:** Among 1093 live births, 99 deaths were recorded between birth and 14 years of follow-up, alongside 44 stillbirths and 10 spontaneous abortions. The majority of deaths occurred within the first six years of life: 22 (22.2%) in the neonatal period, 34 (34.3%) between 28 days and 12 months, and 37 (37.4%) between one and six years, with only 6 (6.1%) deaths between six and 14 years. Respiratory distress was the predominant cause of neonatal death (81%), with half of those babies having low birthweight (<2500 g). Beyond 28 days, malaria became the leading cause, accounting for 45% of deaths between 28 days and 14 years. In adjusted analyses, having a previous live birth was associated with reduced overall mortality (RR 0.52, 95% CI 0.29, 0.94) while low birthweight (<2500 grams) was associated with elevated risk (RR 2.32, 95% CI 1.44, 3.73).

**Discussion and Conclusions:** Child mortality in this semi-rural Beninese cohort was concentrated in early life and driven primarily by respiratory distress in the neonatal period and malaria thereafter. These findings underscore the need for strengthened neonatal care infrastructure, expanded preterm birth support, and scaled-up malaria prevention strategies targeting young children — priorities critical for advancing progress toward SDG 3.2 in West Africa.

**Funding:** This study was funded by the European Developing Countries Clinical Trials Partnership (EDCTP; IP.2007.31080.002) and by the Fondation de France (grant number 00060746 2015). The funders of the study had no role in the study design, data collection or analysis, decision to publish, or preparation of the manuscript.

## Introduction

The United Nations Children’s Fund (UNICEF) defines the under-five mortality rate as the probability that a newborn will die before reaching the age of five years, expressed per 1000 live births [1]. Despite substantial global progress over recent decades, an estimated 9.7 million children under the age of five still die each year worldwide, with approximately 41% of these deaths occurring in sub-Saharan Africa [2]. Under five mortality is driven by a complex interplay of factors including limited access to quality healthcare, infectious diseases, malnutrition, and adverse socio-economic circumstances, with a large proportion of deaths attributable to preventable conditions such as diarrhea, pneumonia, malaria, and HIV [3].

Neonatal mortality is another key indicator of child survival and is defined as death occurring within the first 28 days of life. Known risk factors include advanced maternal age, low socio-economic status, preterm birth, low birth weight, and anemia during pregnancy, while antenatal care uptake and facility-based delivery have been found to be protective factors against neonatal mortality [4]. However, much of this evidence derives from high-income countries or large South and East African cohorts. Prospective longitudinal evidence from West Africa, where epidemiological, environmental, and health system conditions differ substantially, remains limited [4,5]. The only large prospective study in the region, the MOMA study, followed over 20,000 pregnant women across six West African countries (Burkina Faso, Ivory Coast, Mali, Mauritania, Niger and Senegal) but restricted follow-up to eight months postpartum [6], leaving child mortality beyond this period poorly characterized.

In Benin specifically, the under-five mortality rate was estimated at 78 per 1000 live births in 2023 [7]— well above the Sustainable Development Goals (SDG) 3.2 target of 25 — and neonatal mortality at 30 per 1000 live births [7], compared to an SDG target of fewer than 12 [8]. Despite this burden, the evidence base remains critically limited. Existing evidence is restricted either to hospital-based retrospective studies [9,10], national survey data [11], or studies dating from the late 1980s and early 1990s [12,13] which cannot capture community-level deaths, out-of-facility mortality, or survival beyond the neonatal period. The sole community-based effort, a surveillance study using 155 community health workers in the Tanquiéta district between 2006 and 2010 [11], was geographically restricted and did not follow children longitudinally from birth. No prospective community-based cohort with longitudinal follow-up through childhood has previously been conducted in Benin.

To address these gaps, we draw on data from a prospective longitudinal cohort in Benin, with follow-up from pregnancy through adolescence. This study provides a comprehensive descriptive analysis of child mortality in this cohort, including mortality rates, causes of death, and patterns across developmental periods and pregnancy types, and lay the groundwork for future research on child survival in Benin and in West Africa.

## Methods

### Study Population and Original Trial

Data originate from the Malaria in Pregnancy Preventive Alternative Drugs (MiPPAD) clinical trial (ClinicalTrials.gov: NCT00811421), an open-label, randomized controlled trial that recruited 4749 pregnant women between September 1, 2009 and December 1, 2012 during their second trimester across four sub-Saharan African countries [14]. The trial compared the efficacy of two intermittent preventive treatments for malaria in pregnancy (IPTp): sulfadoxine–pyrimethamine (SP) and mefloquine (MQ), administered during the second trimester (≥13 weeks), with a second dose given one month after the first. Each participant also received a long-lasting insecticide-treated bed net (LLIN), that was replaced if damaged through the one-year follow-up.

The present study is an observational follow-up of children born to mothers enrolled in the Benin MiPPAD cohort (N = 1183). Eligible participants were pregnant women at <28 weeks of gestation who were HIV-negative, had no history of allergy to SP or MQ, no history of severe renal, hepatic, psychiatric, or neurological disease, and had not received MQ or halofantrine in the preceding four weeks. All assessments were conducted across three primary health centers serving predominantly semi-rural populations in the Allada district of Benin (Sékou, Allada, and Attogon), approximately 50 km north of Cotonou.

### Maternal Monitoring

Women were monitored throughout pregnancy and during the first year following birth through scheduled and unscheduled visits. During the unscheduled visits, in cases of fever or suspected malaria symptoms (headache, pallor, joint pain), thick blood smear and hemoglobin measurement were performed. Malaria episodes were treated in accordance with national guidelines. Adverse events were systematically recorded throughout follow-up.

### Child Cohort Follow-Up

Longitudinal follow-up data for the child cohort was collected through MiPPAD and two subsequent cohort studies. Within MiPPAD, children were followed up at one month (n = 942), nine months (n = 889), and one year (n = 587) post-birth [15], with mothers also able to bring their infants for unscheduled visits at any time during the first year if the child had a fever or other adverse symptoms. The cohort was subsequently followed up through two ancillary studies: the Exposure to Lead and Manganese and Child Health Risks (EXPLORE) study at six years post-birth (2016–2018; n = 587) [16], and the Child Neurocognitive Development from Birth to Fourteen Years in Benin (DEVINE) study at 13–14 years post-birth (2024–2025; n = 465) [17].

At each MiPPAD visit, assessments included anthropometrics, psychomotor development, and malaria symptoms. Suspected malaria cases were confirmed by thick blood smear and hemoglobin measurement. All unscheduled and emergency visits were provided free of charge. Children with confirmed malaria received artemether-lumefantrine or parenteral treatment in severe cases; anemia was treated with iron syrup or blood transfusion if severe. All treatments were provided at no cost.

### Outcomes

Mortality was examined across four age intervals corresponding to study follow-up waves: birth to one month, one to 12 months, 12 months to six years, and six years to 14 years. For children with an exact date of death recorded (n = 84), age at death was calculated directly and used to derive standard mortality indicators: neonatal mortality (death within the first 28 days), infant mortality (death before one year of age), and under-6 mortality (death before six years of age). For children without an exact date of death but recorded as deceased at a given follow-up wave, death was assigned to the corresponding age interval based on wave timing. For example, a child recorded as deceased at the nine-month visit but with no exact date was classified as having died between one and 12 months. All children that died in the neonatal period had an exact date of death.

Cause-of-death questionnaires were administered retrospectively by interviewers at two time points: the six-year follow-up (EXPLORE, 2016–2018) and the 13–14-year follow-up (DEVINE, 2024–2025), capturing parent-reported causes of death for all children who had died since birth. The questionnaire covered malaria, diarrheal disease, respiratory distress, infections, congenital malformations, genetic disorders, jaundice, and accidental trauma. As cause-of-death data were collected retrospectively at the six- and 13–14-year follow-ups, information was unavailable for 27 of 99 deaths either due to loss to follow-up prior to either visit or parental non-disclosure. Where multiple causes were recorded for a single child, a hierarchical classification was applied based on WHO cause-of-death categories, prioritizing malaria, followed by diarrheal disease, respiratory distress, infection, jaundice, genetic disease, congenital malformation, and trauma or accident [18]. All possible combinations of reported causes of death are presented in **Supplementary Table 1**.

### Covariates

We assessed associations between maternal and birth-related characteristics and mortality outcomes. Explanatory variables included maternal age (years), maternal literacy (ability to read or write; yes/no), previous live birth (yes/no), maternal pre-pregnancy BMI (estimated by subtracting assumed gestational weight gain from weight recorded at inclusion, where weight gain was assumed to be one kg per month after 12 weeks of gestation; pre-pregnancy BMI was then calculated as estimated pre-pregnancy weight divided by height squared [19]), maternal anemia at inclusion (hemoglobin <11 g/dL), maternal anemia at birth (hemoglobin <11 g/dL), preterm birth (<37 gestational weeks; yes/no), low birthweight (<2500 g; yes/no), place of delivery (study health facility vs. other, including home deliveries or non-study health facilities), mode of delivery (caesarean vs. vaginal), multiple birth (yes/no), and child’s sex (male/female).

### Ethical approval

Ethical approval for all studies referenced in this article was granted by the Institutional Review Boards of the Beninese Ethical Committee of the Faculté des Sciences de la Santé (FSS), the Committee of Ethical Research of the Institut des Sciences Biomédicales Appliquées (CER-ISBA) in Benin, and the Consultative Ethics Committee of the Research Institute for Development (IRD) in France. Written informed consent was obtained from all participants, with thumbprints provided by those unable to read or write. Neonates were enrolled in the MiPPAD child cohort with separate consent.

### Statistical analyses

Maternal characteristics were first compared by birth outcome (live birth, stillbirth, spontaneous abortion, and loss to follow-up). Maternal, child, and birth characteristics were then compared between children who died and those who remained under follow-up either at the 6-year and 14-year follow-up periods because not all children participated in both waves. Cause of death was parent-reported for 72 children, and its distribution was examined across four periods: neonatal period, 28 days to 12 months, 12 months to six years, and six years to 14 years. Cause of death and birth characteristics among multiple births were additionally explored.

Mortality incidence rates were calculated using person-time methods across four windows: neonatal (<28 days), infant (<1 year), under-6 years, and overall follow-up to 14 years. Under-6 mortality was reported in place of the conventional under-5 indicator as the study’s follow-up interval spanned one to six years and could not be cleanly split at five years. Exact dates of death were available for 84 of 99 deaths; for the remaining 15, deaths were counted as events where the interval indicator fell completely within the time window. Children without a confirmed death were censored at last known contact. Rates are expressed per 1000 child-days for the neonatal window and per 1000 child-years for all other windows, with exact Poisson 95% confidence intervals calculated using the Garwood method.

Kaplan-Meier survival curves were additionally stratified by prematurity and low birthweight, with children lacking an exact date of death censored at last contact [20]. Risk factors for stillbirths, neonatal mortality, under-one mortality, under-six mortality, and overall mortality (up to 14 years) were assessed using modified Poisson regression with a log link and cluster-robust standard errors at the maternal level to account for non-independence among siblings from multiple births, reported as unadjusted risk ratios (RRs) with 95% confidence intervals. An adjusted modified Poisson regression model was subsequently constructed to evaluate independent predictors of overall mortality which included variables that had a p-value ≤0.25 in the unadjusted overall results. Due to collinearity between preterm birth and low birthweight, low birthweight was retained in the adjusted model because gestational age was assessed using symphysis-fundal height measurement, a field-based method that is less precise than ultrasound dating, whereas birthweight is measured directly and is more accurate and comparable across study sites. All analyses were restricted to observed data.

## Results

A total of 1183 mothers were enrolled in the MiPPAD study. During follow-up until birth, 26 mothers migrated out of the study area, 30 withdrew consent, 14 were lost to follow-up, one did not adhere to the study protocol, and one pregnancy was subsequently determined to be a false pregnancy. Among the remaining pregnancies, 10 mothers experienced a spontaneous abortion (one of which was voluntary) and 42 in stillbirth — including two twin pregnancies, for a total of 44 stillborn infants. In total, there were 1093 live births, including 60 twin births and six triplet births (**Figure 1**). Attendance declined across follow-up waves: 942 children attended the one-month postpartum visit, 889 the nine-month postpartum visit, 874 the 12-month postpartum visit, 587 the six-year assessment, and 468 in the 14-year assessment (**Figure 1**).

**Figure 1:**
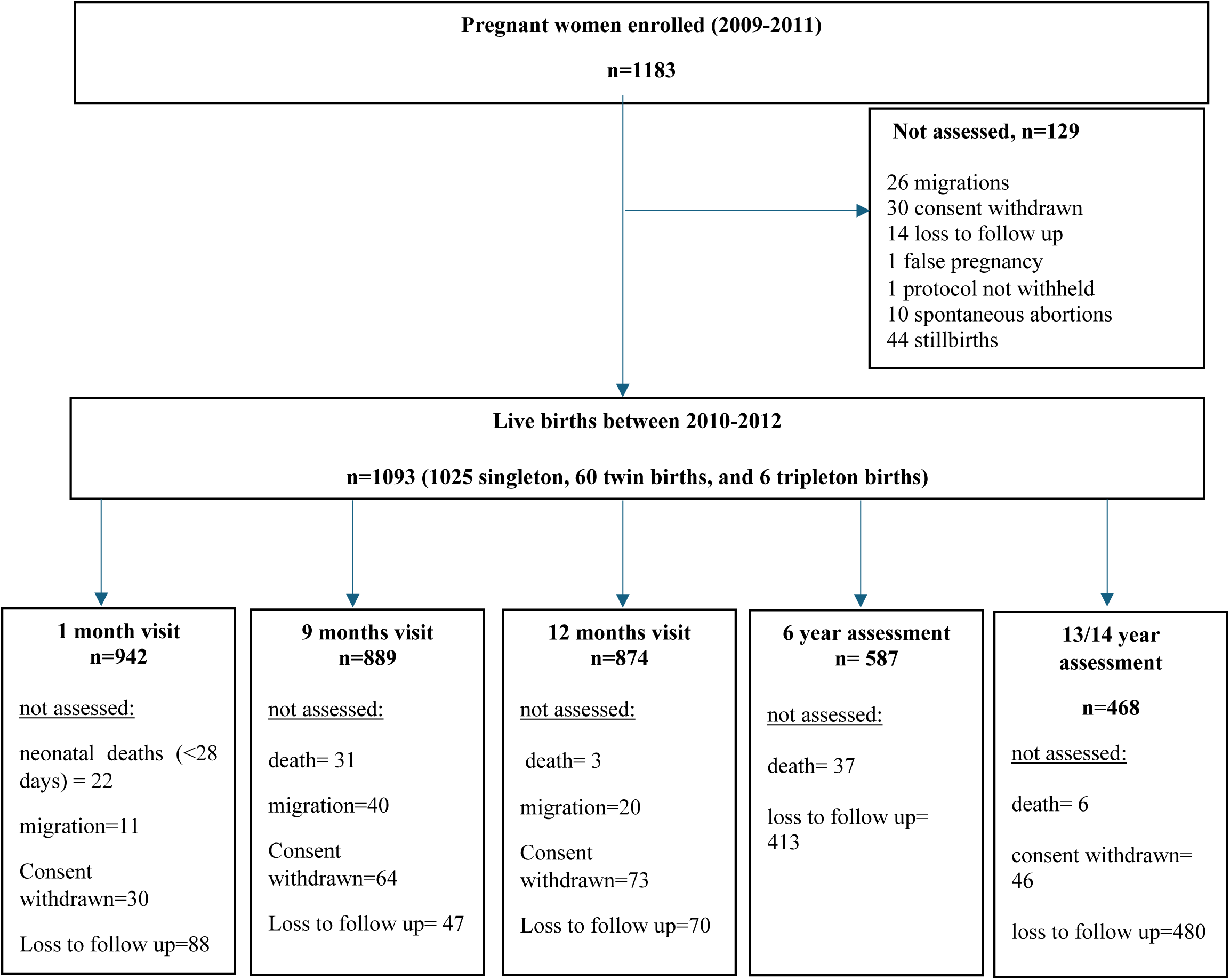
Flow diagram of mothers and their children from the MiPPAD trial and follow-up assessments (2009-2025) Mother-child assessments at 1, 9, and 12 months and at 6 and 13/14 years were independent; attendance at one visit was not required for participation at another. Reasons for non-attendance are reported per visit and do not represent permanent exclusion, except for death. The deaths recording in each wave represent the additional deaths that occurred, not including the previous deaths in the other waves. Multiple births were not included in the 6 and 13/14 year assessment.

The mean gestational age at delivery was 39.5 weeks for live births, 36.1 weeks for stillbirths, and 14.4 weeks for spontaneous abortions (**Table 1**). Among all live births, 23% of mothers were literate, 76% had at least one previous birth, and 93% of deliveries were vaginal. Mothers who experienced a spontaneous abortion were slightly older on average (29.1 years) compared to those with a live birth (25.8 years) or stillbirth (25.8 years) and had a higher rate of illiteracy (40%). Among mothers with stillbirth, the majority had no prior stillbirth (90%) or spontaneous abortion (71%) and most carried a singleton pregnancy (91%)(**Table 1**). Compared to mothers with a live birth, those lost to follow-up during pregnancy were slightly more educated (32% vs. 23% literacy) and had a prior spontaneous abortion (30% vs. 20%) (**Table 1**).

**Table 1:** Maternal characteristics by birth status.

| Variables | Live Birth<br>N = 1093<sup>1</sup> | Still Birth<br>N = 44<sup>1</sup> | Spontaneous Abortion<br>N = 10<sup>1</sup> | Loss to Follow Up<br>N = 72<sup>1</sup> |
| --- | --- | --- | --- | --- |
| **Maternal age at inclusion (years)** | 25.8 (5.5) | 25.8 (6.3) | 29.1 (8.5) | 24.2 (4.8) |
| **Maternal literacy (can read or write)** | 246 (23%) | 6 (14%) | 4 (40%) | 23 (32%) |
| *Missing* | 0 | 0 | 0 | 1 |
| **Number of living children** |  |  |  |  |
| 0 | 263 (24%) | 19 (43%) | 4 (40%) | 24 (34%) |
| 1 | 215 (20%) | 7 (16%) | 2 (20%) | 17 (24%) |
| 2-3 | 394 (37%) | 9 (20%) | 2 (20%) | 22 (31%) |
| ≥4 | 206 (19%) | 9 (20%) | 2 (20%) | 8 (11%) |
| *Missing* | 15 | 0 | 0 | 1 |
| **Pre-pregnancy BMI (kg/m2)** | 21.5 (3.5) | 22.2 (5.1) | 24.5 (6.9) | 21.7 (3.0) |
| *Missing* | 1 | 1 | 0 | 0 |
| **Mid-upper arm circumference (cm)** | 25.4 (2.7) | 25.2 (3.3) | 26.5 (5.2) | 25.3 (2.3) |
| *Missing* | 0 | 0 | 0 | 1 |
| **Gestational age at inclusion (weeks)** | 21.9 (4.2) | 21.3 (4.1) | 11.5 (4.0) | 21.4 (4.9) |
| **Anaemia at inclusion (Hemoglobin <11 g/dL)** | 744 (68%) | 26 (59%) | 6 (60%) | 46 (64%) |
| **Previous live birth (No)** | 234 (22%) | 15 (37%) | 3 (30%) | 21 (30%) |
| *Missing* | 13 | 3 | 0 | 1 |
| **Previous spontaneous abortions (n)** |  |  |  |  |
| 0 | 863 (79.8%) | 29 (71%) | 7 (70%) | 49 (69%) |
| 1 | 172 (16%) | 8 (20%) | 1 (10%) | 20 (28%) |
| 2 | 37 (3.4%) | 4 (9%) | 2 (20%) | 1 (1%) |
| 3 | 7 (0.6%) | 0 (0%) | 0 (0%) | 1 (1%) |
| 4 | 2 (0.2%) | 0 (0%) | 0 (0%) | 0 (0%) |
| *Missing* | 12 | 3 | 0 | 1 |
| **Previous stillbirths (n)** |  |  |  |  |
| 0 | 1027 (95%) | 37 (90%) | 10 (100%) | 69 (97%) |
| 1 | 48 (4.5%) | 2 (5%) | 0 (0%) | 2 (3%) |
| 2 | 3 (0.5%) | 2 (5%) | 0 (0%) | 0 (0%) |
| *Missing* | 15 | 3 | 0 | 1 |
| **Gestational age at pregnancy outcome (weeks)** | 39.5 (2.2) | 36.1 (4.9) | 14.4 (3.0) | – |
| *Missing* | 30 | 4 | 1 | 72 |
| **Plurality** |  |  |  |  |
| *Singleton* | 1027 (94%) | 40 (91%) | 3 (100%) | – |
| *Twin* | 60 (5.5%) | 4 (9%) | 0 (0%) | – |
| *Triplet* | 6 (0.5%) | 0 (0%) | 0 (0%) | – |
| *Missing* | 0 | 0 | 7 | 72 |

<sup>1</sup>Median (Q1, Q3); n (%)

Of the 99 deaths occurring between birth and 14 years of follow-up, the majority occurred within the first six years of life: 22 in the neonatal period (including three on the day of birth), 34 between 28 days and 12 months, and 37 between one and six years. Only six deaths were recorded between six and 14 years. Compared with children followed up at six or 14 years, those who died in the neonatal period had a markedly higher prevalence of very low birthweight (29% vs 0.3%), low birthweight (38% vs 10.3%), and preterm birth, including very preterm (9% vs 0.2%), moderately preterm (27% vs 0.9%), and late preterm (28 to <37 weeks) (14% vs 5.1%). However, one child born extremely preterm (<28 weeks) survived to 14 years of age. Neonatal deaths were also more frequently associated with delivery outside the study facility (41% vs 15%) and caesarean delivery (27% vs 5%) (**Table 2**). For deaths occurring between 1 and 6 years, mothers had the lowest rate of illiteracy (8%) compared to child deaths previous or after as well as were more nulliparous (39%) compared to those alive and followed up at 6 or 14 years (17%). For pre-term births, the majority happened between 34 and less than 37 weeks. A higher percentage of deaths among multiple births occurred during the neonatal period compared to any other period. The majority of child deaths were boys in almost every death interval (neonatal (67%); between 28 days and under one (56%); and between 6 and 14 years (83%)) except for deaths between one and six which had a slightly higher incidence of death among girls (59%) (**Table 2**). Children who were lost to follow-up for both the six- and 14-year waves differed from those who participated in either the 6- or 14-year follow-up: they had higher maternal literacy rates (28% vs. 21%), a slightly higher proportion of low birthweight (18% vs. 10%), and a higher proportion of multiple births (11% vs. 3%) (**Table 2**).

**Table 2:** Maternal, birth, and child characteristics by timing of death and follow-up status up to 14 years.

| Variable | Live<br>births<br>N=1093<sup>1</sup> | Neonatal<br>death<br>(<28<br>days)<br>N = 22<sup>1</sup> | Post-neonatal<br>death (28<br>days–<1<br>year)<br>N = 34<sup>1</sup> | Death<br>between<br>1–6<br>years<br>N = 37<sup>1</sup> | Death<br>between<br>6–14<br>years<br>N = 6<sup>1</sup> | Lost to<br>follow-up<br>at 6 and 14<br>years<br>N = 327<sup>1</sup> | Alive,<br>followed<br>up at 6 or<br>14 years<br>N = 667<sup>1</sup> | Total<br>deaths<br>N=99<sup>1</sup> |
| --- | --- | --- | --- | --- | --- | --- | --- | --- |
| **Maternal age**<br>**(years)** | 25.0<br>(21.0,<br>30.0) | 26.5<br>(20.0,<br>30.0) | 27.5 (20.0,<br>32.0) | 24.0<br>(20.0,<br>25.0) | 27.0<br>(20.0,<br>28.0) | 25.0 (20.0,<br>30.0) | 25.0 (22.0,<br>30.0) | 25.0<br>(20.0,<br>30.0) |
| **Maternal**<br>**literacy (any)** | 246<br>(23%) | 5 (23%) | 8 (24%) | 3 (8.1%) | 1 (17%) | 92 (28%) | 137 (21%) | 17 (17%) |
| **Number of**<br>**living children** |  |  |  |  |  |  |  |  |
| *0* | 278<br>(25%) | 6 (27%) | 10 (29%) | 17<br>(46%) | 2 (33%) | 98 (30%) | 145 (22%) | 35 (35%) |
| *1* | 215<br>(20%) | 3 (14%) | 8 (24%) | 9 (24%) | 0 (0%) | 64 (20%) | 131 (20%) | 20 (20%) |
| *2-3* | 394<br>(36%) | 9 (41%) | 9 (26%) | 10<br>(27%) | 3 (50%) | 112 (34%) | 251 (38%) | 31 (31%) |
| *≥4* | 206<br>(19%) | 4 (18%) | 7 (21%) | 1 (3%) | 1 (17%) | 53 (16%) | 140 (21%) | 13 (13%) |
| **Previous live**<br>**birth (No)** | 234<br>(22%) | 6 (27%) | 10 (29%) | 14<br>(39%) | 2 (33%) | 88 (27%) | 114 (17%) | 32 (33%) |
| *Missing* | 13 | 0 | 0 | 1 | 0 | 3 | 9 | 1 |
| **Maternal**<br>**weight at**<br>**inclusion (kg)** | 55.0<br>(50.0,<br>61.0) | 52.8<br>(47.5,<br>55.9) | 55.8 (50.5,<br>62.5) | 53.0<br>(48.5,<br>58.0) | 50.8<br>(50.0,<br>55.0) | 55.0 (51.0,<br>62.0) | 55.0 (50.0,<br>61.0) | 53.0<br>(48.5,<br>58.0) |
| **Pre-pregnancy**<br>**BMI (kg/m<sup>2</sup>)** | 21.0<br>(19.4,<br>23.0) | 21.1<br>(20.1,<br>22.3) | 21.0 (19.5,<br>23.2) | 20.2<br>(19.1,<br>21.4) | 20.1<br>(19.7,<br>20.2) | 21.5 (19.7,<br>23.5) | 20.9 (19.2,<br>22.8) | 20.4<br>(19.3,<br>22.0) |
| *Missing* | 1 | 0 | 0 | 1 | 0 | 0 | 0 | 1 |
| **Anaemia at**<br>**inclusion**<br>**(Hemoglobin**<br>**<11 g/dL)** | 744<br>(68%) | 18 (82%) | 28 (82%) | 23<br>(62%) | 3 (50%) | 236 (72%) | 436 (65%) | 72 (73%) |
| **Pre-term birth**<br>**categories** |  |  |  |  |  |  |  |  |
| *Extremely*<br>*preterm (<28*<br>*weeks)* | 1 (0.1%) | 0 (0%) | 0 (0%) | 0 (0%) | 0 (0%) | 0 (0%) | 1 (0.2%) | 0 (0%) |
| *Very preterm*<br>*(28 to <32*<br>*weeks)* | 5 (0.5%) | 2 (9%) | 0 (0%) | 0 (0%) | 0 (0%) | 2 (0.6%) | 1 (0.2%) | 2 (2.1%) |
| *Moderately*<br>*preterm (32 to*<br>*<34 weeks)* | 18<br>(1.7%) | 6 (27%) | 0 (0%) | 1 (3%) | 0 (0%) | 5 (1.6%) | 6 (0.9%) | 7 (7.2%) |
| *Late preterm*<br>*(34 to <37*<br>*weeks)* | 60<br>(5.6%) | 3 (14%) | 1 (3%) | 3 (8%) | 1 (17%) | 18 (5.8%) | 34 (5.1%) | 8 (8.2%) |
| *Term (≥37*<br>*weeks)* | 979<br>(92.1%) | 11 (50%) | 32 (97%) | 32<br>(89%) | 5 (83%) | 286 (92%) | 613<br>(93.6%) | 80<br>(82.5%) |
| *Missing* | 30 | 0 | 1 | 1 | 0 | 16 | 12 | 2 |

| **Birthweight category** |  |  |  |  |  |  |  |  |
| --- | --- | --- | --- | --- | --- | --- | --- | --- |
| *Very low birthweight (<1500g)* | 9 (0.8%) | 6 (29%) | 0 (0%) | 0 (0%) | 0 (0%) | 1 (0.3%) | 2 (0.3%) | 6 (6%) |
| *Low birthweight (1500–<2500g)* | 147 (13.6%) | 8 (38%) | 11 (32%) | 6 (16%) | 1 (17%) | 53 (16.4%) | 68 (10.3%) | 26 (27%) |
| *Normal birthweight (≥2500g)* | 928 (85.6%) | 7 (33%) | 23 (68%) | 31 (84%) | 5 (83%) | 269 (83.3%) | 593 (89.4%) | 66 (67%) |
| *Missing* | 9 | 1 | 0 | 0 | 0 | 4 | 4 | 1 |
| **Anaemia at birth (Hemoglobin <11 g/dL)** | 391 (39%) | 7 (32%) | 13 (45%) | 15 (41%) | 1 (17%) | 131 (48%) | 224 (36%) | 36 (38%) |
| *Missing* | 101 | 0 | 5 | 0 | 0 | 54 | 42 | 5 |
| **Place of delivery (Study health facility)** | 871 (80%) | 13 (59%) | 28 (82%) | 33 (89%) | 5 (83%) | 229 (70%) | 563 (85%) | 79 (80%) |
| *Missing* | 1 | 0 | 0 | 0 | 0 | 0 | 1 |  |
| **Mode of delivery (Vaginal)** | 1021 (93%) | 16 (73%) | 32 (94%) | 35 (95%) | 6 (100%) | 297 (91%) | 635 (95%) | 89 (90%) |
| **Plurality** |  |  |  |  |  |  |  |  |
| *Singleton* | 1027 (94%) | 14 (64%) | 28 (82%) | 37 (100%) | 6 (100%) | 295 (90.5%) | 647 (97%) | 85 (86%) |
| *Twin* | 60 (5.5%) | 7 (32%) | 6 (18%) | 0 (0%) | 0 (0%) | 31 (9.5%) | 16 (2.4%) | 13 (13%) |
| *Triplet* | 6 (0.5%) | 1 (4%) | 0 (0%) | 0 (0%) | 0 (0%) | 1 (0.3%) | 4 (0.6%) | 1 (1%) |
| **Child's sex (Female)** | 551 (51%) | 7 (33%) | 15 (44%) | 22 (59%) | 1 (17%) | 167 (51%) | 339 (51%) | 45 (46%) |
| *Missing* | 3 | 1 | 0 | 0 | 0 | 1 | 1 | 1 |

<sup>1</sup>Median (Q1, Q3); n (%)

Birth characteristics of multiple births and cause of death are presented in **Supplementary Table 2**. Among multiple births who died during the neonatal period, 38% were preterm and all were low birthweight, compared with 17% and 65%, respectively, among multiple births who were alive and followed up at 6 or 14 years. No deaths were recorded between 1 and 14 years among multiple births.

Of the 99 recorded deaths, cause of death information was parent-reported for 72 children (**Table 3**). During the neonatal period, respiratory distress was the predominant cause of death (n=13, 81%), and half of the recorded deaths for respiratory distress were underweight babies (<2500 grams). Between 28 days and 12 months, malaria was the most reported cause (n=8, 32%), followed by respiratory distress (n=7, 28%) and infection (n=6, 24%). Between one and six years, malaria accounted for the majority of deaths (n=21, 81%) as well as for deaths between six and 12 years (n=3, 75%). For children who had malaria as cause of death before one year, the majority of them (5 out of 8) happened after six months post-partum. Among children from multiple births with recorded death (n = 14), a total of 9 had recorded cause of death. Respiratory distress was the leading cause between birth and one year (78%, n = 7), with one death attributed to malaria and one to infection (**Supplementary Table 2**).

**Table 3:** Parent-reported causes of child death, N=72.

|  | Neonatal death<br>(<28 days)<br>N = 22<sup>1</sup> | Infant death<br>(28 days– <1<br>year)<br>N = 34<sup>1</sup> | Death between<br>1–6 years<br>N = 37<sup>1</sup> | Death<br>between<br>6–14 years<br>N = 6<sup>1</sup> | Total<sup>1</sup><br>N= 99 |
| --- | --- | --- | --- | --- | --- |
| **Cause of death** |  |  |  |  |  |
| Malaria | 0 (0%) | 8 (32%) | 21 (81%) | 3 (75%) | 32 (45%) |
| Diarrhea | 0 (0%) | 3 (12%) | 1 (3.8%) | 1 (25%) | 5 (7%) |
| Respiratory distress | 13 (81%) | 7 (28%) | 0 (0%) | 0 (0%) | 20 (28%) |
| Infection | 0 (0%) | 6 (24%) | 1 (3.8%) | 0 (0%) | 7 (10%) |
| Jaundice | 3 (19%) | 0 (0%) | 0 (0%) | 0 (0%) | 3 (4.2%) |
| Genetic disease | 0 (0%) | 0 (0%) | 1 (3.8%) | 0 (0%) | 1 (1.4%) |
| Congenital malformation | 0 (0%) | 1 (4%) | 0 (0%) | 0 (0%) | 1 (1.4%) |
| Trauma / accident | 0 (0%) | 0 (0%) | 2 (7.6%) | 0 (0%) | 2 (3%) |
| Missing | 6 | 9 | 11 | 2 | 28 |

<sup>1</sup>n (%)

### Mortality rates in person-time

For neonatal mortality, 22 deaths occurred over 30 050 child-days, yielding a mortality rate of 0.73 per 1000 child-days (95% CI: 0.46, 1.11). For infant mortality, 56 deaths occurred over 996 child-years, yielding a rate of 56.2 per 1000 child-years (95% CI: 42.5, 73.0). Under-6 mortality comprised 93 deaths over 4549 child-years (20.4 per 1000 child-years; 95% CI: 16.5, 25.0). Over the entire follow-up period, 99 deaths occurred over 7886 child-years, yielding an overall mortality rate of 12.6 per 1000 child-years (95% CI: 10.2, 15.3).

### Kaplan Meier curves

**Figure 2** displays Kaplan-Meier survival curves stratified by birthweight category (low birthweight <2500g vs normal birthweight ≥2500g). Of the 1093 children in the sample, birthweight information was available for 1039 (95%), of whom 889 had normal birthweight and 150 had low birthweight. Survival probability declined steeply in the low birthweight group during the first 1000 days of life compared to a more gradual decline in the normal birthweight group (p < 0.0001). After approximately 1000 days, both curves plateaued, suggesting that low birthweight children who survived early life faced a mortality risk more comparable to their normal birthweight peers.

**Figure 2:**
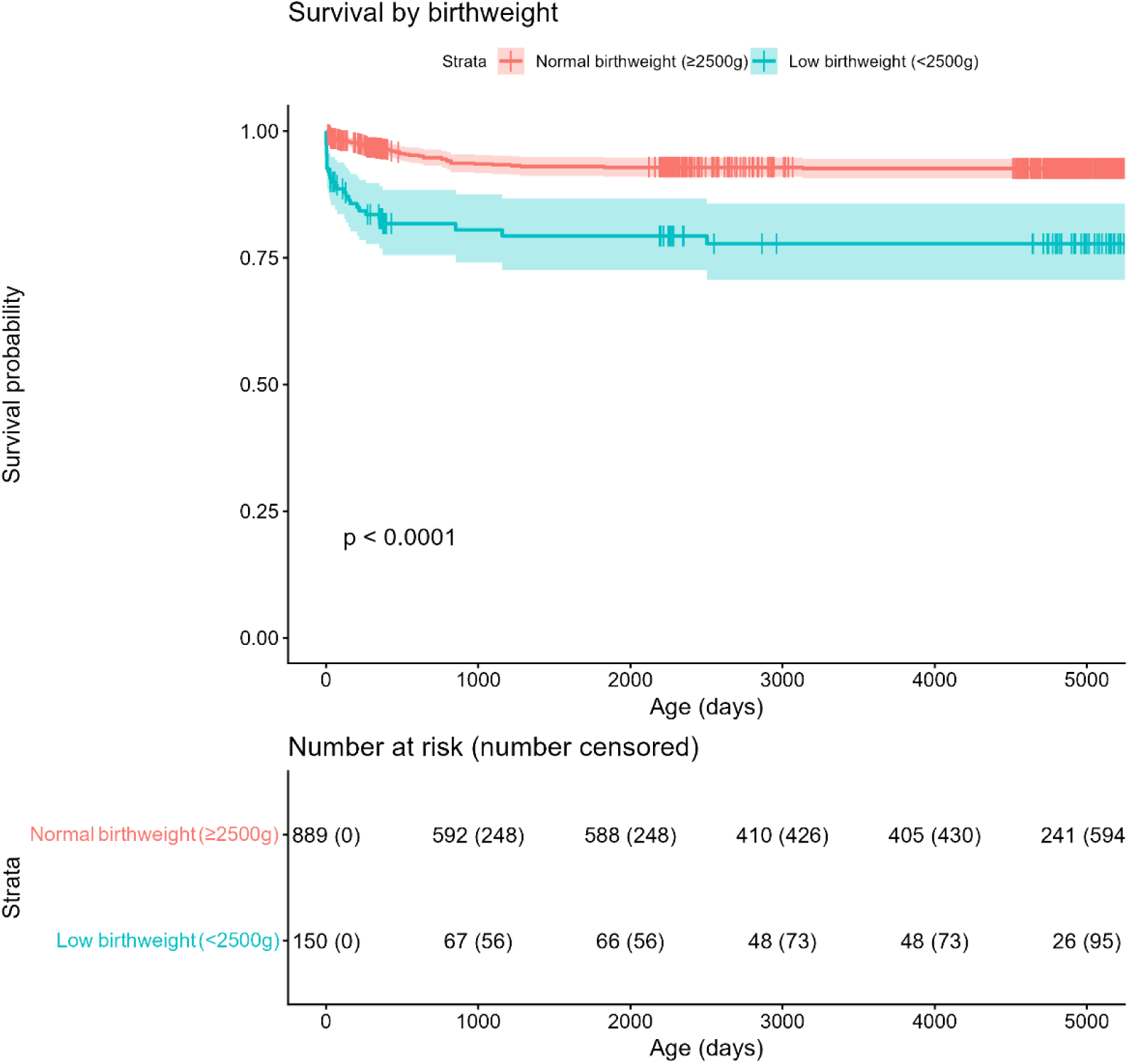
Kaplan-Meier survival curves by birthweight category (low birthweight <2500g vs normal birthweight ≥2500g) from birth to approximately 14 years of age. Shaded areas represent 95% confidence intervals. Vertical tick marks on the curves indicate censored observations (children who were lost to follow-up or did not experience the event by the end of the study period). The number at risk table below the figure shows the number of children remaining under follow-up at each 1000-day interval as well as the number of children censored at each interval. P-value derived from the log-rank test.

A similar pattern was observed by gestational age (**Figure 3**). Of the 1093 children, gestational age data were available for 1023 (93%), of whom 82 were born prematurely (<37 weeks) and 941 at term. Premature births showed a steep decline in survival probability during the first 1000 days compared to term births (p < 0.0001), with the gap between groups narrowing considerably thereafter, suggesting that, as with low birthweight, the excess mortality risk associated with prematurity was concentrated in early life.

**Figure 3:**
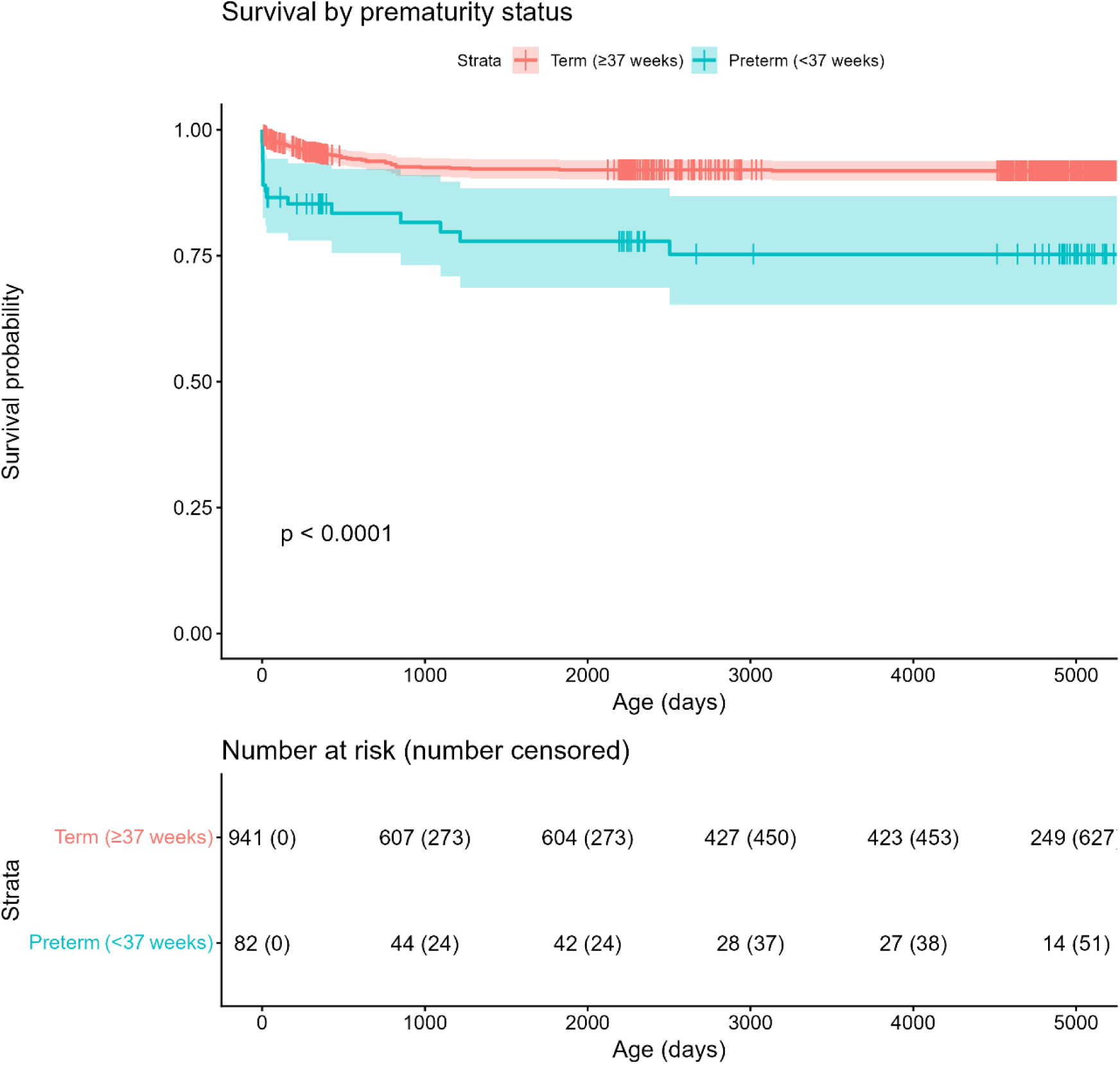
Kaplan-Meier survival curves by gestational age category (premature birth <37 weeks vs term birth ≥37 weeks) from birth to approximately 14 years of age. Shaded areas represent 95% confidence intervals. Vertical tick marks on the curves indicate censored observations (children who were lost to follow-up or did not experience the event by the end of the study period). The number at risk table below the figure shows the number of children remaining under follow-up at each 1000-day interval as well as the number of children censored at each interval. P-value derived from the log-rank test.

### Unadjusted risk ratios

In unadjusted analyses (**Supplementary Table 3**), having a previous live birth was associated with a lower risk of stillbirth (RR 0.49, 95% CI 0.25, 0.96).

For neonatal mortality, preterm birth (RR 11.65, 95% CI 4.89, 27.77), low birthweight (RR 11.90, 95% CI 4.72, 30.01), and multiple birth (RR 8.89, 95% CI 3.51, 22.55) were all strongly associated with elevated risk. Caesarean delivery (RR 5.32, 95% CI 1.98, 14.25) and delivery outside the study facility (RR 2.73, 95% CI 1.10, 6.74) were also associated with increased neonatal mortality.

Similar patterns were observed for under-one mortality, though effect sizes were attenuated: preterm birth (RR 3.25, 95% CI 1.73, 6.10), low birthweight (RR 4.96, 95% CI 2.93, 8.38), multiple birth (RR 5.19, 95% CI 2.85, 9.46), and caesarean delivery (RR 2.36, 95% CI 1.05, 5.34). Additionally, anemia at inclusion (hemoglobin <11 g/dL) was associated with elevated risk of child mortality (RR 2.16, 95% CI 1.10, 4.25).

For under-six mortality, having a previous live birth was associated with reduced risk (RR 0.57, 95% CI 0.38, 0.87), while higher pre-pregnancy BMI was associated with a modest increase in risk (RR 1.01, 95% CI 1.00, 1.02). Preterm birth (RR 2.49, 95% CI 1.50, 4.13), low birthweight (RR 3.02, 95% CI 2.00, 4.57), and multiple birth (RR 2.76, 95% CI 1.57, 4.85) remained significantly associated with mortality.

For overall mortality up to 14 years, findings were consistent with earlier periods. Having a previous live birth was associated with a decreased risk in mortality (RR 0.58, 95% CI 0.39, 0.87) whereas pre-pregnancy BMI (RR 1.01, 95% CI 1.00, 1.02), preterm birth (RR 2.45, 95% CI 1.50, 3.98), low birthweight (RR 2.84, 95% CI 1.90, 4.24), and multiple birth (RR 2.53, 95% CI 1.45, 4.44) were associated with a higher risk in mortality (**Supplementary Table 3**).

### Adjusted risk ratios

For overall mortality up to 14 years, we adjusted for maternal age, literacy status, parity, pre-pregnancy BMI, low birthweight, mode of delivery, and multiple birth. After adjustment, having a previous live birth remained associated with reduced risk of child mortality (RR 0.52, 95% CI 0.29, 0.94), while low birthweight (RR 2.32, 95% CI 1.44, 3.73) remained associated with elevated risk (**Table 4**).

**Table 4:** Adjusted overall death Poisson regression results, N=1070.

| Variables | RR (95% CI) | P value |
| --- | --- | --- |
| **Overall deaths (up to 14 years) (n=100)** |  |  |
| *Maternal age at inclusion* | 1.01 (0.97, 1.07) | 0.55 |
| *Mother literate (yes)* | 0.69 (0.40, 1.18) | 0.17 |
| *Previous live birth (Yes)* | 0.52 (0.29, 0.94) | 0.03* |
| *Pre-pregnancy BMI* | 0.97 (0.90, 1.04) | 0.38 |
| *Low birthweight (<2500 grams)* | 2.32 (1.44, 3.73) | <0.001*** |
| *Mode of delivery (Caesarean)* | 1.22 (0.62, 2.40) | 0.56 |
| *Multiple birth (yes)* | 1.43 (0.77, 2.64) | 0.25 |

The table presents the adjusted risk ratios (RRs) with 95% confidence intervals (CIs) and p-values estimated using modified Poisson regression models with a log link and cluster-robust standard errors at the maternal level. Only mothers with complete data on the covariates of interest were used for the analysis.
RR: Risk Ratios 95% CI: 95% Confidence Intervals
\* $P < 0.05$
\*\* $P \leq 0.01$
\*\*\* $P \leq 0.001$

## Discussion

This study summarizes the distributions and trends of child mortality from birth to 14 years in a longitudinal birth cohort in semi-rural Benin where mothers took antimalarials throughout pregnancy and were offered a LLIN to use throughout pregnancy up to the first year after birth. Of 1093 live births, 99 deaths were observed across childhood, with the majority occurring before age six, consistent with existing literature showing that most child deaths occur under age five and are attributable to treatable infectious diseases and preterm and low birth weight birth complications [21]. Overall, malaria was the leading cause of death across childhood (n=32, 45%), which is unsurprising given Benin’s exceptionally high malaria burden. Benin represents one of the highest incidences of malaria in the world [22,23], and throughout the study period, incidence remained persistently high, ranging from approximately 364 to 444 per 1000 population (2009-2018) and tapering off to 354 per 1000 in 2024.

Among neonatal deaths, respiratory distress was the most commonly parent-reported cause of death. Half of these neonates had low birthweight (<2500 grams) and half were preterm, with the majority born between 32 and less than 34 weeks of gestation. This pattern is consistent with the broader literature, in which respiratory distress syndrome is the leading cause of death among preterm and low-birthweight neonates in sub-Saharan Africa, a region that accounts for approximately 80% of neonatal deaths globally [24]. In our analyses, both low birthweight and preterm birth were associated with higher mortality risk in unadjusted models; after adjustment, low birthweight remained independently associated with increased risk of mortality through age 14. This is unsurprising given that these infants face compounding vulnerabilities from birth, including impaired respiration, feeding difficulties, poor thermoregulation, and elevated infection risk as well as a risk for later developmental disabilities or chronic conditions compared to normal birth weight children [25,26].

In this study, neonatal deaths represent the one window in childhood where malaria was not the dominant cause, reflecting the protective role of maternal immunity in early infancy. Newborns benefit from maternal IgG antibodies, fetal hemoglobin, and breastfeeding, all of which inhibit parasite growth [27]. As these protections wane around six to nine months of age, children become increasingly vulnerable to malaria. This is reflected in our findings, where malaria was the leading cause of death (n=32, 45%), with the majority of those deaths occurring after 6 months postpartum (n=29), suggesting that tapering maternal antibodies left these infants at greater risk. This pattern continued and intensified through early childhood. Among deaths occurring between 1 and 6 years, malaria accounted for 81% (n=26) of deaths, consistent with previous research identifying children aged one to four as most susceptible to malaria in high-endemic settings [27]. A similar pattern was observed among the 6-to-14-year age group, where malaria remained the primary cause of death, though the smaller number of deaths in this window warrants cautious interpretation.

Multiple births were associated with elevated mortality risk across all periods in the unadjusted models, but this association attenuated after adjusting for low birthweight. This pattern suggests that low birthweight may partly explain the elevated mortality risk observed among multiple births, possibly acting as an intermediary on the causal pathway, though this analysis was not designed to formally assess mediation. Of the 14 deaths occurring among multiple births, the majority took place in the neonatal period (n=8), with five attributed to respiratory distress as the primary cause. This aligns with existing literature highlighting several contributing factors, including poorer management of multiple pregnancies, a higher likelihood of birth defects, increased obstetric risk relative to singleton pregnancies, and greater susceptibility to growth retardation, preterm birth, and delivery complications [28].

Among maternal factors, having a previous live birth was consistently associated with reduced mortality risk across the unadjusted analyses, including lower risk of stillbirth and child mortality overall. This is consistent with previous literature reporting higher birthweights and better weight gain during infancy among infants born to multiparous compared to nulliparous women [29]. Additionally, primiparous women are known to be at higher risk of prenatal malaria, which has in turn been associated with increased rates of preterm birth and low birthweight, both strong predictors of child mortality [30,31]. Although we were unable to assess prenatal malaria exposure directly in this cohort, this pathway may partly explain the elevated mortality risk observed among children of primiparous women.

In unadjusted analyses, higher pre-pregnancy BMI was associated with a small increase in mortality risk, and maternal anemia at inclusion was associated with increased under-one mortality — findings that, while attenuated in the adjusted model, point to the broader role of maternal nutritional status in shaping child survival [32]. The association between caesarean delivery and neonatal and under-one mortality in unadjusted analyses most likely reflects indication bias, whereby caesarean delivery is performed in the context of pre-existing obstetric complications rather than being a direct cause of mortality, though we did not have information on pregnancy or birth complications.

These results provide evidence that more services are needed to help pre-term, low birthweight children, and that after one year, more services are needed to address malaria risk among children who no longer are protected by the antibodies of their mothers. Additionally, in a world with growing antibiotics, strategies to protect children against malaria while also preventing antibiotic resistance are needed.

### Implications

#### Malaria prevention during pregnancy

Benin has since strengthened its malaria prevention in pregnancy protocol in ways that directly address the gaps observed in this cohort. Women in this study received two doses of IPTp-SP or IPTp-MQ rather than the now-recommended three, which likely contributed to the high rates of malaria during pregnancy and placental malaria observed here [14], and in turn, to the elevated preterm birth and neonatal mortality we report [31,33]. Current guidelines mandate three doses, and distribution has been extended to community health workers, improving coverage in semi-rural and rural areas where access to formal care is most limited [34]. Completing three doses has been associated with a birthweight increase of more than 0.16 kg, consistent with a dose-dependent relationship between IPTp and birthweight [35], and improved adherence combined with greater community-level access represents a concrete pathway through which the outcomes observed here could be reduced under current practice.

Additionally, the use of LLIN by pregnant mothers can help reduce malaria during pregnancy. Bed net coverage among pregnant women in Benin is high, largely attributable to a 2017 mass distribution campaign, but the uptake of nets provided at ANC visits remains poor. One study in Benin found while around 63% of women received an LLIN at their first visit, only about 11% were using it, with most continuing to rely on older, damaged nets [36]. These findings suggest that distribution alone is insufficient. Pairing it with routine net quality checks, active encouragement of consistent use during pregnancy, and targeted education, particularly in the private sector where distribution has lagged, could meaningfully reduce malaria in pregnancy.

#### Malaria prevention in early childhood

In 2023, malaria affected 39% of children under five in Benin, with a mortality rate of approximately 106 deaths per 100,000 children [37]. Responding to this burden, Benin has built a layered prevention strategy combining vaccination, chemoprevention, and vector control. On 25 April 2024, Benin introduced the RTS,S/AS01 vaccine for children, administering four doses starting at five months of age across 16 of 34 health districts [38]. Although vaccine efficacy against infection alone is modest (∼36%) [39], one real-world analysis found that the vaccine alone prevents roughly one in eight child deaths attributable to malaria [40]. Additionally, its impact is substantially greater in combination with other tools. When the vaccine is paired with bed nets and chemoprevention, efficacy in reducing clinical malaria rises above 90% [38].

In addition to the vaccine, Seasonal Malaria Chemoprevention (SMP) and Perennial Malaria Chemoprevention (PMC) are two methods to reduce malaria infection in infants and children. SMP includes providing children ages 3 months to 59 months with monthly SP plus amodiaquine over three to four cycles during peak malaria transmission [41]. In Benin, as of 2023, SMP has reached 15 communes across six health zones, with 82% coverage of the target population [41]. In parallel, Benin participates in a Unitaid-supported PMC programme that provides infants (from birth up to 24 months) with a minimum of eight SP treatments timed to the immunization schedule [42].

Despite these advances, important gaps remain. Vaccine access is currently limited to 16 of 34 health zones, with full national extension expected by 2027, and eligibility beyond 18 months deserves consideration given that a substantial proportion of malaria deaths occur in children aged one to six years. Sustaining impact will ultimately depend on closing the financing and implementation gaps that continue to limit reach in the most underserved communities.

### Neonatal care for premature and low-birthweight infants

Given that the majority of neonatal deaths occurred among premature and low-birthweight infants for whom respiratory distress was the leading reported cause of death, targeted infrastructure investment is urgently needed. Preventing respiratory distress syndrome mortality requires a continuum of care spanning antenatal corticosteroids for lung maturation, safe birth management, and immediate neonatal resuscitation — steps that are feasible even in low-resource settings. Because few newborns need advanced resuscitation, training birth attendants in basic resuscitation alone can avert many intrapartum-related deaths. Thermal care strategies such as kangaroo mother care and early initiation of breastmilk, which protects against necrotizing enterocolitis, are similarly low-cost and effective. Where gaps remain, blended oxygen and CPAP devices can prevent hypoxia but are largely inaccessible in resource-limited environments, underscoring the need for affordable, context-adapted equipment paired with trained personnel. Low birthweight, as a readily measurable field indicator, represents a practical entry point for identifying at-risk neonates and triggering timely intervention [24,43].

### Limitations and Strengths

This study has some limitations that must be acknowledged. First, cause of death was reported by parents rather than determined through verbal autopsy, introducing the possibility of cause of death misclassification. Additionally, not all parents were willing to disclose death information to field workers. Second, while the official date of death was unavailable for 15 of the 99 recorded deaths, the longitudinal nature of the study allowed us to classify timing of death based on asking the parents at the time of each study wave if their child was living or not. For neonatal deaths, we were able to obtain the exact date of death, so the 15 unrecorded dates of death occurred between 28 days and 6 years. Third, substantial loss to follow-up occurred at both the 6-year and 14-year time points. For this reason, we report mortality using person-time observation rather than per 1000 births. Fourth, given that preterm births were more prevalent among those lost to follow-up, it is likely that additional deaths went unrecorded in this group, meaning our mortality estimates may understate the true rate. Fifth, prenatal malaria could not be assessed for the full cohort, as malaria testing was restricted to mothers enrolled in a nested sub-study on anemia and malaria.

Despite these limitations, this study has several important strengths. To our knowledge, this is the first cohort of its kind in West Africa to follow children and their mothers from pregnancy through 14 years of age — a follow-up period that remains ongoing. The quality of our mortality data is also a notable strength: for nearly all recorded deaths, we captured both the exact timing and cause of death, which represents a clear advantage over DHS data, which often restricts mortality timing to predefined recall periods and may lack cause-of-death information altogether. The prospective nature of the study further minimizes recall bias and allows for more accurate ascertainment of exposures and outcomes compared to retrospective approaches, including the direct assessment of prematurity and low birthweight — two critical determinants of early child survival that are frequently underreported in routine health data. By focusing on a semi-rural population, this study also fills an important gap in the literature, as prior research in Benin has largely been based on hospital records from urban settings, limiting generalizability to the broader population. Finally, the data generated provide current, population-level mortality estimates that can directly inform public health interventions, particularly those targeting neonatal deaths and malaria-related mortality.

## Conclusion

This study represents one of the first longitudinal birth cohorts in Benin to examine child mortality in a semi-rural district. Over 14 years of follow-up, 99 deaths were recorded — though the extent of loss to follow-up suggests the true burden of child mortality may be considerably higher. The leading causes of death were malaria and respiratory distress, the latter occurring predominantly among preterm infants, highlighting the particular vulnerability of this population in the neonatal period. These findings underscore the need for targeted interventions, including strengthening neonatal care infrastructures, expanding access to preterm birth support, and scaling up infectious disease prevention strategies for young children. Addressing these priorities will be essential for reducing child mortality in Benin and advancing progress toward the SDG target of ending preventable deaths in children under five.

## Data Availability

Deidentified individual data and the data dictionary will be made available to others upon reasonable requests to the project leader, Florence Bodeau-Livinec at. A data transfer agreement will be signed between EHESP and the requesting institution before data sharing.

## Acknowledgments

We would like to acknowledge and sincerely thank the local healthcare providers in Allada, Benin who participated in all studies mentioned within this article.

## Author contributions

K.M.B. contributed to study design, data collection and analysis, interpretation of results, and manuscript writing. M.C. contributed to data management and manuscript writing. M.J.A. contributed to interpretation of results and manuscript writing. R.Z. contributed to data collection and manuscript writing. C.F. contributed to data management. A.M. contributed to data collection and manuscript writing. F.B.L. contributed to study design, interpretation of results, and manuscript writing.

All authors had full access to all data in the study and accept responsibility for the decision to submit for publication. K.M.B., M.C., R.Z., C.F. and F.B.L. directly accessed the underlying data reported in this manuscript at the level of the individual participant; the accuracy and integrity of the data were independently verified by M.C., R.Z., C.F., and F.B.L.

## Supplementary Information

**Table 1:** children recorded as multiple causes of death, n=72.

| **Multiple causes of death (n=72)** | **N (%)** |
| --- | --- |
| Malaria + Infection | 23 (49%) |
| Infection + Respiratory distress | 7 (15%) |
| Diarrhea + Infection | 4 (8%) |
| Jaundice + Malaria + Infection | 3 (6%) |
| Congenital malformation + Respiratory distress | 2 (4%) |
| Diarrhea + Malaria | 2 (4%) |
| Diarrhea + Malaria + Infection | 2 (4%) |
| Diarrhea + Jaundice + Malaria + Infection | 1 (2%) |
| Diarrhea + Jaundice + Infection | 1 (2%) |
| Jaundice + Malaria | 1 (2%) |
| Jaundice + Respiratory distress | 1 (2%) |

For parents who reported multiple causes of death for their child, all possible combinations are presented.

**Table 2:** Birth characteristics and cause of death among multiple births, n=66.

For parents who reported multiple causes of death for their child, all possible combinations are presented.
| Variables | Live multiple births<sup>1</sup><br>N=66 | Neonatal death (<28 days)<br>N = 8<sup>1</sup> | Post-neonatal death (28 days–<1 year)<br>N = 6<sup>1</sup> | Lost to follow-up at 6 and 14 years<br>N = 32<sup>1</sup> | Alive, followed up at 6 or 14 years<br>N = 20<sup>1</sup> | Total deaths<sup>1</sup><br>N=14 |
| --- | --- | --- | --- | --- | --- | --- |
| **Maternal age (years)** | 27.0 (23.0, 32.0) | 30.0 (23.5, 30.0) | 25.0 (21.0, 28.0) | 29.0 (23.5, 34.0) | 26.0 (24.0, 30.0) | 28.0 (21.0, 30.0) |
| **Maternal literacy (any)** | 10 (15%) | 1 (13%) | 3 (50%) | 4 (13%) | 2 (10%) | 4 (29%) |
| **Number of living children** |  |  |  |  |  |  |
| 0 | 10 (15%) | 2 (25%) | 2 (33%) | 3 (9.4%) | 3 (15%) | 4 (29%) |
| 1 | 12 (18%) | 0 (0%) | 3 (50%) | 4 (13%) | 5 (25%) | 3 (21%) |
| 2-3 | 30 (45%) | 6 (75%) | 1 (17%) | 16 (50%) | 7 (35%) | 7 (50%) |
| ≥4 | 14 (21%) | 0 (0%) | 0 (0%) | 9 (28%) | 5 (25%) | 0 (0%) |
| **Previous live birth (No)** | 10 (16%) | 2 (25%) | 2 (33%) | 3 (10%) | 3 (15%) | 4 (29%) |
| **Missing** | 2 | 0 | 0 | 59.0 (53.0, 64.8) | 52.0 (47.5, 58.0) |  |
| **Pre-pregnancy BMI (kg/m<sup>2</sup>)** | 21.6 (20.1, 22.9) | 20.9 (20.1, 22.8) | 20.8 (18.4, 21.8) | 22.6 (21.3, 24.3) | 20.9 (19.5, 21.6) | 20.9 (20.1, 22.8) |
| **Anaemia at inclusion (hemoglobin <11 g/dL)** | 55 (83%) | 7 (88%) | 6 (100%) | 28 (88%) | 14 (70%) | 13 (93%) |
| **Gestational age at delivery** |  |  |  |  |  |  |
| *Extremely preterm (<28 weeks)* | 0 (0%) | 0 (0%) | 0 (0%) | 0 (0%) | 0 (0%) | 0 (0%) |
| *Very preterm (28 to <32 weeks)* | 0 (0%) | 0 (0%) | 0 (0%) | 0 (0%) | 0 (0%) | 0 (0%) |
| *Moderately preterm (32 to <34 weeks)* | 4 (6%) | 3 (38%) | 0 (0%) | 1 (3%) | 0 (0%) | 3 (21%) |
| *Late preterm (34 to <37 weeks)* | 8 (13%) | 0 (0%) | 1 (17%) | 4 (13%) | 3 (17%) | 1 (7%) |
| *Term (≥37 weeks)* | 51 (81%) | 5 (63%) | 5 (83%) | 26 (84%) | 15 (83%) | 10 (71%) |
| **Missing** | 3 | 0 | 0 | 1 | 2 |  |
| **Birthweight** |  |  |  |  |  |  |
| *Very low birthweight (<1500g)* | 9 (14%) | 6 (86%) | 0 (0%) | 1 (3%) | 2 (10%) | 6 (46%) |
| *Low birthweight (1500–<2500g)* | 39 (60%) | 1 (14%) | 5 (83%) | 22 (69%) | 11 (55%) | 6 (46%) |
| *Normal birthweight (≥2500g)* | 17 (26%) | 0 (0%) | 1 (17%) | 9 (28%) | 7 (35%) | 1 (8%) |
| **Missing** | 1 | 1 | 0 | 0 | 0 | 1 |
| **Anaemia at birth (hemoglobin <11 g/dL)** | 27 (47%) | 3 (38%) | 1 (25%) | 18 (64%) | 5 (29%) | 4 (33%) |
| **Missing** | 9 | 0 | 2 | 4 | 3 | 2 |
| **Place of delivery (study health facility)** | 39 (59%) | 3 (38%) | 3 (50%) | 19 (59%) | 14 (70%) | 6 (43%) |
| **Mode of delivery (Vaginal)** | 53 (80%) | 3 (38%) | 5 (83%) | 27 (84%) | 18 (90%) | 8 (57%) |
| **Plurality** |  |  |  |  |  |  |
| *Singleton* | 0 (0%) | 0 (0%) | 0 (0%) | 0 (0%) | 0 (0%) | 0 (0%) |
| *Twin* | 60 (89.9%) | 7 (88%) | 6 (100%) | 31 (97%) | 16 (80%) | 13 (93%) |
| *Triplet* | 6 (9.1%) | 1 (13%) | 0 (0%) | 1 (3%) | 4 (20%) | 1 (7%) |
| **Child's sex (Female)** | 28 (43%) | 4 (57%) | 1 (17%) | 11 (34%) | 12 (60%) | 5 (38%) |
| **Missing** | 1 | 1 | 0 | 0 | 0 | 1 |
| **Cause of death** |  |  |  |  |  |  |
| *Malaria* | - | 0 (0%) | 1 (25%) | - | - | 1 (11%) |
| *Diarrhea* | - | 0 (0%) | 0 (0%) | - | - | 0 (0%) |
| *Respiratory distress* | - | 5 (100%) | 2 (50%) | - | - | 7 (78%) |
| *Infection* | - | 0 (0%) | 1 (25%) | - | - | 1 (11%) |
| *Jaundice* | - | 0 (0%) | 0 (0%) | - | - | 0 (0%) |
| *Genetic disease* | - | 0 (0%) | 0 (0%) | - | - | 0 (0%) |
| *Congintal malformation* | - | 0 (0%) | 0 (0%) | - | - | 0 (0%) |
| *Trauma/Accident* | - | 0 (0%) | 0 (0%) | - | - | 0 (0%) |
| *Missing* | - | 3 | 2 | - | - | 5 |

**Table 3:** Unadjusted risk ratios for neonatal, under-one-year, and under-six mortality according to maternal, child, and birth characteristics.

| Outcome (n= number of deaths) | Variables | RR (95% CI) | P value | N of observations |
| --- | --- | --- | --- | --- |
| **Stillbirths (n=44)** |  |  |  |  |
|  | *Maternal age at inclusion* | 1.00 (0.94, 1.06) | 0.98 | 1137 |
|  | *Mother literate(yes)* | 0.55 (0.21, 1.46) | 0.23 | 1137 |
|  | *Previous live birth (Yes)* | 0.49 (0.25, 0.96) | 0.04* | 1121 |
|  | *Pre-pregnancy BMI* | 1.04 (0.96, 1.13) | 0.34 | 1135 |
|  | *Anemia (hemoglobin <11 g/dL)* | 0.73 (0.36, 1.50) | 0.39 | 1137 |
| **Neonatal (<28 days) (n=22)** |  |  |  |  |
|  | *Maternal age at inclusion* | 1.00 (0.92, 1.08) | 0.91 | 1093 |
|  | *Mother literate(yes)* | 1.01 (0.37, 2.78) | 0.98 | 1093 |
|  | *Previous live birth (Yes)* | 0.74 (0.28, 1.91) | 0.53 | 1092 |
|  | *Pre-pregnancy BMI* | 0.97 (0.88, 1.07) | 0.53 | 1093 |
|  | *Anemia at inclusion (hemoglobin <11 g/dL)* | 2.11 (0.71, 6.30) | 0.18 | 1093 |
|  | *Anemia at birth(hemoglobin <11 g/dL)* | 0.72 (0.27, 1.92) | 0.51 | 992 |
|  | *Preterm (<37 weeks)* | 11.65 (4.89, 27.77) | <0.001*** | 1063 |
|  | *Low birthweight (<2500g)* | 11.90 (4.72, 30.01) | <0.001*** | 1084 |
|  | *Delivery location (other)* | 2.73 (1.10, 6.74) | 0.03 | 1092 |
|  | *Mode of delivery (Caesarean)* | 5.32 (1.98, 14.25) | <0.001*** | 1093 |
|  | *Multiple birth (yes)* | 8.89 (3.51, 22.55) | <0.001*** | 1093 |
|  | *Child's sex (Male)* | 2.04 (0.75, 5.56) | 0.16 | 1090 |
| **Under-1 year (n=56)** |  |  |  |  |
|  | *Maternal age at inclusion* | 1.01 (0.97, 1.06) | 0.57 | 1093 |
|  | *Mother literate(yes)* | 1.04 (0.55, 1.97) | 0.90 | 1093 |
|  | *Previous live birth (Yes)* | 0.69 (0.38, 1.24) | 0.22 | 1080 |
|  | *Pre-pregnancy BMI* | 1.01 (0.93, 1.09) | 0.96 | 1093 |
|  | *Anemia at inclusion (hemoglobin <11 g/dL)* | 2.16 (1.10, 4.25) | 0.03* | 1093 |
|  | *Anemia at birth (hemoglobin <11 g/dL)* | 0.99 (0.56, 1.76) | 0.98 | 992 |
|  | *Preterm (<37 weeks)* | 3.25 (1.73, 6.10) | <0.001*** | 1063 |
|  | *Low birthweight (<2500g)* | 4.96 (2.93, 8.38) | <0.001*** | 1084 |
|  | *Delivery location (other)* | 1.44 (0.78, 2.67) | 0.25 | 1092 |
|  | *Mode of delivery (Caesarean)* | 2.36 (1.05, 5.34) | 0.04* | 1093 |
|  | *Multiple birth (yes)* | 5.19 (2.85, 9.46) | <0.001*** | 1093 |
|  | *Child's sex (Male)* | 1.53 (0.90, 2.62) | 0.12 | 1090 |
| **Under-6 (n=93)** |  |  |  |  |
|  | *Maternal age at inclusion* | 0.98 (0.94, 1.02) | 0.28 | 1093 |
|  | *Mother literate(yes)* | 0.72 (0.42, 1.23) | 0.23 | 1093 |
|  | *Previous live birth (Yes)* | 0.57 (0.38, 0.87) | 0.01* | 1080 |
|  | *Pre-pregnancy BMI* | 1.01 (1.00, 1.02) | 0.003** | 1093 |
|  | *Anemia at inclusion (hemoglobin <11 g/dL)* | 1.35 (0.86, 2.11) | 0.19 | 1093 |
|  | *Anemia at birth (hemoglobin <11 g/dL)* | 1.02 (0.67, 1.54) | 0.94 | 992 |
|  | *Preterm (<37 weeks)* | 2.49 (1.50, 4.13) | <0.001*** | 1063 |
|  | *Low birthweight (<2500g)* | 3.02 (2.00, 4.57) | <0.001*** | 1084 |
|  | *Delivery location (other)* | 1.01 (0.61, 1.69) | 0.96 | 1092 |
|  | *Mode of delivery (Caesarean)* | 1.71 (0.86, 3.38) | 0.12 | 1093 |
|  | *Multiple birth (yes)* | 2.76 (1.57, 4.85) | <0.001*** | 1093 |
|  | *Child's sex (Male)* | 1.12 (0.75, 1.65) | 0.59 | 1090 |
| **Overall (to 14 years) (n=100)** |  |  |  |  |
|  | *Maternal age at inclusion* | 0.98 (0.94, 1.02) | 0.25 | 1093 |
|  | *Mother literate(yes)* | 0.71 (0.42, 1.19) | 0.19 | 1093 |
|  | *Previous live birth (Yes)* | 0.58 (0.39, 0.87) | 0.01* | 1080 |
|  | *Pre-pregnancy BMI* | 1.01 (1.00, 1.02) | 0.04* | 1093 |
|  | *Anemia at inclusion (hemoglobin <11 g/dL)* | 1.25 (0.82, 1.92) | 0.30 | 1093 |
|  | *Anemia at birth (hemoglobin <11 g/dL)* | 0.98 (0.66, 1.46) | 0.92 | 992 |
|  | *Preterm (<37 weeks)* | 2.45 (1.50, 3.98) | <0.001*** | 1063 |
|  | *Low birthweight (<2500g)* | 2.84 (1.90, 4.24) | <0.001*** | 1084 |
|  | *Delivery location (other)* | 0.99 (0.60, 1.62) | 0.95 | 1092 |
|  | *Mode of delivery (Caesarean)* | 1.58 (0.80, 3.11) | 0.19 | 1093 |
|  | *Multiple birth (yes)* | 2.53 (1.45, 4.44) | 0.001** | 1093 |
|  | *Child's sex (Male)* | 1.20 (0.82, 1.76) | 0.28 | 1090 |

The table presents the unadjusted risk ratios (RRs) with 95% confidence intervals (CIs) and p-values estimated using modified Poisson regression models with a log link and cluster-robust standard errors at the maternal level.
RR: Risk Ratios 95% CI: 95% Confidence Intervals
\* P< 0.05
\*\* P ≤ 0.01
\*\*\* P ≤ 0.001

## References

[1] UNICEF. Child Mortality. UNICEF DATA n.d. https://data.unicef.org/topic/child-survival/under-five-mortality/ (accessed January 12, 2026).

[2] Rutherford ME, Mulholland K, Hill PC. How access to health care relates to under-five mortality in sub-Saharan Africa: systematic review. Trop Med Int Health 2010;15:508–19. 10.1111/j.1365-3156.2010.02497.x.

[3] Avelino IC, Van-Dúnem J, Varandas L. Under-five mortality and social determinants in africa: a systematic review. Eur J Pediatr 2025;184:150. 10.1007/s00431-024-05966-w.

[4] Kefale B, Jancey J, Gebremedhin AT, Belay DG, Nyadanu SD, Pereira G, et al. Risk factors for neonatal mortality: an umbrella review of systematic reviews and meta-analyses. eClinicalMedicine 2025;88. 10.1016/j.eclinm.2025.103525.

[5] Campbell A, Rudan I. Systematic review of birth cohort studies in Africa. J Glob Health 2011;1:46–58.

[6] Chalumeau M, Salanave B, Bouvier-Colle MH, de Bernis L, Prual A, Bréart G. Risk factors for perinatal mortality in West Africa: a population-based study of 20326 pregnancies. MOMA group. Acta Paediatr 2000;89:1115–21. 10.1080/713794568.

[7] World Bank Open Data. World Bank Open Data n.d. https://data.worldbank.org/indicator/SH.DYN.MORT (accessed January 12, 2026).

[8] WHO. End preventable deaths of newborns and children under 5 years of age n.d. https://www.who.int/data/gho/data/themes/topics/sdg-target-3_2-newborn-and-child-mortality (accessed January 12, 2026).

[9] Houssou M, Hessou YGA, Sawadogo B, Antara S, Mckenzie A, Sawadogo M, et al. Neonatal mortality and risk factors in the University Hospital of the Mother and Child Lagoon in Cotonou, Benin, 2015-2016. J Interv Epidemiol Public Health 2020;3.

[10] Kounou A, Koudokpon H, Sintondji K, Lègba B, Fabiyi K, Yadouléton A, et al. Prevalence and determinants of neonatal infections in Benin based on a retrospective study in six reference hospitals. Sci Rep 2025;15:11093. 10.1038/s41598-025-94442-y.

[11] Imourou BCA, Perini P, Sohoundé L, Ahanhanzo C. [Community-based surveillance of maternal, infant, and child (under-5) mortality in the health district of Tanguiéta (Benin) from 2006 through 2010]. Med Sante Trop 2013;23:332–6. 10.1684/mst.2013.0240.

[12] Velema JP, Alihonou EM, Chippaux JP, van Boxel Y, Gbedji E, Adegbini R. Malaria morbidity and mortality in children under three years of age on the coast of Benin, West Africa. Trans R Soc Trop Med Hyg 1991;85:430–5. 10.1016/0035-9203(91)90206-e.

[13] Tomlinson AJ, Healey CH, Koetsier MA. Perinatal mortality in northern Benin. Trop Doct 1997;27:172–3. 10.1177/004947559702700320.

[14] González R, Mombo-Ngoma G, Ouédraogo S, Kakolwa MA, Abdulla S, Accrombessi M, et al. Intermittent Preventive Treatment of Malaria in Pregnancy with Mefloquine in HIV-Negative Women: A Multicentre Randomized Controlled Trial. PLoS Med 2014;11:e1001733. 10.1371/journal.pmed.1001733.

[15] Bodeau-Livinec F, Zoumenou RE, Fermanian C, Massougbodji A. The MiPPAD birth cohort of Benin - Follow-up at one year 2026. 10.23708/ZCIPAX.

[16] Bodeau-Livinec F, Zoumenou RE, Fermanian C, Massougbodji A. The MiPPAD birth cohort of Benin - Follow-up at six years 2026. 10.23708/WZ9A8P.

[17] Bodeau-Livinec F, Zoumenou RE, Fermanian C, Massougbodji A. The MiPPAD birth cohort of Benin - Follow-up at fourteen years 2026. 10.23708/LZCJLC.

[18] WHO. End preventable deaths of newborns and children under 5 years of age n.d. https://www.who.int/data/gho/data/themes/topics/sdg-target-3_2-newborn-and-child-mortality (accessed March 6, 2026).

[19] Ouédraogo S, Koura GK, Accrombessi MMK, Bodeau-Livinec F, Massougbodji A, Cot M. Maternal anemia at first antenatal visit: prevalence and risk factors in a malaria-endemic area in Benin. Am J Trop Med Hyg 2012;87:418–24. 10.4269/ajtmh.2012.11-0706.

[20] Rich JT, Neely JG, Paniello RC, Voelker CCJ, Nussenbaum B, Wang EW. A practical guide to undertanding Kaplan-Meier curves. Otolaryngol--Head Neck Surg Off J Am Acad Otolaryngol-Head Neck Surg 2010;143:331–6. 10.1016/j.otohns.2010.05.007.

[21] Perin J, Mulick A, Yeung D, Villavicencio F, Lopez G, Strong KL, et al. Global, regional, and national causes of under-5 mortality in 2000–19: an updated systematic analysis with implications for the Sustainable Development Goals. Lancet Child Adolesc Health 2022;6:106–15. 10.1016/S2352-4642(21)00311-4.

[22] Charting the Globe. Incidence of malaria per 1,000 population at risk in Benin - ChartingTheGlobe n.d. https://chartingtheglobe.com/region/benin/health/disease-prevalence?indicator=malaria&visual=map (accessed March 12, 2026).

[23] WHO. Malaria incidence (per 1000 population at risk). World Health Organ Data n.d. https://data.who.int/indicators/i/B868307/442CEA8 (accessed March 12, 2026).

[24] Rosa-Mangeret F, Benski A-C, Golaz A, Zala PZ, Kyokan M, Wagner N, et al. 2.5 Million Annual Deaths—Are Neonates in Low- and Middle-Income Countries Too Small to Be Seen? A Bottom-Up Overview on Neonatal Morbi-Mortality. Trop Med Infect Dis 2022;7:64. 10.3390/tropicalmed7050064.

[25] Cao G, Liu J, Liu M. Global, Regional, and National Incidence and Mortality of Neonatal Preterm Birth, 1990-2019. JAMA Pediatr 2022;176:787–96. 10.1001/jamapediatrics.2022.1622.

[26] Stein REK, Siegel MJ, Bauman LJ. Are Children of Moderately Low Birth Weight at Increased Risk for Poor Health? A New Look at an Old Question. Pediatrics 2006;118:217–23. 10.1542/peds.2005-2836.

[27] Ranjha R, Singh K, Baharia RK, Mohan M, Anvikar AR, Bharti PK. Age-specific malaria vulnerability and transmission reservoir among children. Glob Pediatr 2023;6:100085. 10.1016/j.gpeds.2023.100085.

[28] Bhusal MK, Khanal SP. A Systematic Review of Factors Associated with Under-Five Child Mortality. BioMed Res Int 2022;2022:1181409. 10.1155/2022/1181409.

[29] Garces A, Perez W, Harrison MS, Hwang KS, Nolen TL, Goldenberg RL, et al. Association of parity with birthweight and neonatal death in five sites: The Global Network’s Maternal Newborn Health Registry study. Reprod Health 2020;17:182. 10.1186/s12978-020-01025-3.

[30] Guyatt HL, Snow RW. Impact of Malaria during Pregnancy on Low Birth Weight in Sub-Saharan Africa. Clin Microbiol Rev 2004;17:760–9. 10.1128/cmr.17.4.760-769.2004.

[31] Fried M, Kurtis JD, Swihart B, Pond-Tor S, Barry A, Sidibe Y, et al. Systemic Inflammatory Response to Malaria During Pregnancy Is Associated With Pregnancy Loss and Preterm Delivery. Clin Infect Dis 2017;65:1729–35. 10.1093/cid/cix623.

[32] Christian P, Mullany LC, Hurley KM, Katz J, Black RE. Nutrition and maternal, neonatal, and child health. Semin Perinatol 2015;39:361–72. 10.1053/j.semperi.2015.06.009.

[33] Garrison A, Boivin MJ, Fiévet N, Zoumenou R, Alao JM, Massougbodji A, et al. The Effects of Malaria in Pregnancy on Neurocognitive Development in Children at 1 and 6 Years of Age in Benin: A Prospective Mother-Child Cohort. Clin Infect Dis Off Publ Infect Dis Soc Am 2022;74:766–75. 10.1093/cid/ciab569.

[34] WHO. Intermittent preventative treatment to reduce the risk of malaria during pregnancy n.d. https://www.who.int/tools/elena/interventions/iptp-pregnancy (accessed March 16, 2026).

[35] Quakyi I, Tornyigah B, Houze P, Kusi KA, Coleman N, Escriou G, et al. High uptake of Intermittent Preventive Treatment of malaria in pregnancy is associated with improved birth weight among pregnant women in Ghana. Sci Rep 2019;9:19034. 10.1038/s41598-019-55046-5.

[36] Cottrell G, Djènontin A, Soares C, Bouraima A, Fiogbé M, Egbinola S, et al. Suboptimal distribution and utilization of antenatal care given bed Nets undermine pregnant women’s protection in Benin: a prospective field study. BMC Public Health 2025;25:2529. 10.1186/s12889-025-22212-6.

[37] Malaria in Benin: Statistics & Facts | Severe Malaria Observatory n.d. https://www.severemalaria.org/countries/benin (accessed June 1, 2026).

[38] GAVI. Rolling out vaccines to beat malaria together: Time to harness the power of immunisation for a malaria-free future n.d. https://www.gavi.org/news-insights/insight-reports/rolling-out-vaccines-beat-malaria-together-time-harness-power-immunisation-malaria-free-future (accessed June 4, 2026).

[39] WHO. WHO recommends groundbreaking malaria vaccine for children at risk n.d. https://www.who.int/news/item/06-10-2021-who-recommends-groundbreaking-malaria-vaccine-for-children-at-risk (accessed March 16, 2026).

[40] Mwapasa V, Asante KP, Milligan P, Akech S, Oduro A, Mathanga DP, et al. Impact of introducing RTS,S/AS01E malaria vaccine on mortality in young children in Ghana, Kenya, and Malawi: an observational evaluation of a cluster-randomised implementation programme. The Lancet 2026;407:1796–808. 10.1016/S0140-6736(26)00248-5.

[41] WHO guidelines for malaria n.d. https://www.who.int/publications/i/item/guidelines-for-malaria (accessed June 1, 2026).

[42] WHO. Updated WHO recommendations for malaria chemoprevention among children and pregnant women n.d. https://www.who.int/news/item/03-06-2022-Updated-WHO-recommendations-for-malaria-chemoprevention-among-children-and-pregnant-women (accessed March 16, 2026).

[43] Wall SN, Lee AC, Niermeyer S, English M, Keenan WJ, Carlo W, et al. Neonatal resuscitation in low-resource settings: What, who, and how to overcome challenges to scale up? Int J Gynaecol Obstet Off Organ Int Fed Gynaecol Obstet 2009;107:S47–64. 10.1016/j.ijgo.2009.07.013.

